# Clinical Decision Support Tool and Rapid Point-of-Care Platform for Determining Disease Severity in Patients with COVID-19

**DOI:** 10.1101/2020.04.16.20068411

**Authors:** Michael P. McRae, Glennon W. Simmons, Nicolaos J. Christodoulides, Zhibing Lu, Stella K. Kang, David Fenyo, Timothy Alcorn, Isaac P. Dapkins, Iman Sharif, Deniz Vurmaz, Sayli S. Modak, Kritika Srinivasan, Shruti Warhadpande, Ravi Shrivastav, John T. McDevitt

## Abstract

SARS-CoV-2 is the virus that causes coronavirus disease (COVID-19) which has reached pandemic levels resulting in significant morbidity and mortality affecting every inhabited continent. The large number of patients requiring intensive care threatens to overwhelm healthcare systems globally. Likewise, there is a compelling need for a COVID-19 disease severity test to prioritize care and resources for patients at elevated risk of mortality. Here, an integrated point-of-care COVID-19 Severity Score and clinical decision support system is presented using biomarker measurements of C-reactive protein (CRP), N-terminus pro B type natriuretic peptide (NT-proBNP), myoglobin (MYO), D-dimer, procalcitonin (PCT), creatine kinase–myocardial band (CK-MB), and cardiac troponin I (cTnI). The COVID-19 Severity Score combines multiplex biomarker measurements and risk factors in a statistical learning algorithm to predict mortality. The COVID-19 Severity Score was trained and evaluated using data from 160 hospitalized COVID-19 patients from Wuhan, China. Our analysis finds that COVID-19 Severity Scores were significantly higher for the group that died versus the group that was discharged with median (interquartile range) scores of 59 (40–83) and 9 (6–17), respectively, and area under the curve of 0.94 (95% CI 0.89– 0.99). These promising initial models pave the way for a point-of-care COVID-19 Severity Score system to impact patient care after further validation with externally collected clinical data. Clinical decision support tools for COVID-19 have strong potential to empower healthcare providers to save lives by prioritizing critical care in patients at high risk for adverse outcomes.

## Introduction

The 2019-20 pandemic of coronavirus disease 2019 (COVID-19) caused by the severe acute respiratory syndrome coronavirus 2 (SARS-CoV-2)^1^ was first reported in Wuhan, Hubei, China, in December 2019.^2^ On March 11, 2020, the World Health Organization (WHO) declared the outbreak a pandemic.^3^ Although there is expected to be a substantial under-reporting of cases (particularly of persons with milder symptoms, asymptomatic cases, and in countries with low testing volume), as of April 4, 2020 over 1M cases have been confirmed with approximately 60,000 deaths from the disease globally and major outbreaks in the US, Italy, China, and Spain.^4^ Symptoms of COVID-19 are non-specific, and infected individuals may develop fever, cough, fatigue, shortness of breath, or muscle aches with further disease development leading to severe pneumonia, acute respiratory distress syndrome (ARDS), myocardial injury, sepsis, septic shock, and death.^5, 6^ The median incubation period is approximately five days, and 97.5% of those who develop symptoms will do so within 11.5 days.^7^ A larger analysis of 2449 patients reported hospitalization rates of 20 to 31 percent and ICU admission rates of 4.9 to 11.5 percent.^8^ This large number of patients requiring intensive care threatens to overwhelm healthcare systems around the world. There is a need for a COVID-19 disease severity test to prioritize care for patients at elevated risk of mortality and manage low risk patients in outpatient settings or at home through self-quarantine.

Biomarker tests provide key information about the health or disease status of an individual, including COVID-19. In an analysis of 127 hospitalized COVID-19 patients in Wuhan, China, the most common complications leading to death were acute cardiac injury (58.3%), ARDS (55.6%), coagulation dysfunction (38.9%), and acute kidney injury (33.3%).^9^ Biomarkers, such as cardiac troponin I (cTnI), C-reactive protein (CRP), D-dimer, and procalcitonin (PCT) were significantly increased in those that died versus those that recovered with prognostic values (as determined by area under the curve [AUC]) of 0.939, 0.870, 0.866, and 0.900, respectively. In another study, data from 82 COVID-19 deaths found that respiratory, cardiac, hemorrhage, hepatic, and renal damage were present in 100%, 89%, 80.5%, 78.0%, and 31.7% of patients, respectively, in which most patients had increased CRP (100%) and D-dimer (97.1%).^10^ The importance of D-dimer as a prognostic factor was also demonstrated with odds of death significantly increased for levels greater than 1µg/mL on admission.^11^ A biomarker of cardiac failure, N-terminal pro-B-type natriuretic peptide (NT-proBNP) has also been shown to be predictive of death in patients with community acquired pneumonia.^12^ A recent study of 416 hospitalized patients with COVID-19 reported 82 patients (19.7%) had cardiac injury,^13^ in which patients with myocardial damage had significantly higher levels of CRP, PCT, creatine kinase-myocardial band (CK-MB), cTnI, and NT-proBNP. Patients with cardiac injury also more frequently required noninvasive mechanical ventilation (46.3% vs. 3.9%) or invasive mechanical ventilation (22.0% vs. 4.2%) and experienced higher rates of complications such as ARDS (58.5% vs. 14.7%) compared to patients without cardiac injury. Ultimately, patients with cardiac injury had higher mortality than those without it (51.2% vs. 4.5%). Given such data, others have recommended elevating treatment priority and aggressiveness for patients with underlying cardiovascular disease and evidence of cardiac injury.^14^ This growing body of clinical evidence related to COVID-19 disease severity suggests that biomarkers can play a dominant role in a scoring system to identify COVID-19 patients with increased risk of severe disease and mortality.

While there are multiple commercially available platforms for COVID-19 diagnosis based on molecular detection of the viral RNA, there remains a significant gap in determining disease prognosis with respect to early identification of individuals that are at elevated risk of mortality. Identifying and monitoring those at risk of severe complications is critical for both resource planning and prognostication. Likewise, ruling out and/or reducing the admission of patients with very low risk of complications who can be safely managed through self-quarantine would conserve precious medical resources during a surge of new cases in an outbreak. While clinical decision support tools have been developed for sepsis disease severity^15^ and are in development for COVID-19 disease severity,^16^ to our knowledge there are no scoring systems for COVID-19 disease severity that are intricately linked to the biomarker tests at the point of care or based on lab-on-a-chip platforms. Access to an integrated test and scoring system for use at the point of care and in low- and middle-income countries would help to manage this disease on a global basis.

In this study, we describe our most recent work toward developing the programmable bio nano chip (p-BNC) with the capacity to learn^17^ and adapting it to the task of assessing COVID-19 disease severity. This multiplex and multiclass platform has been demonstrated previously for the detection and quantitation of protein biomarkers, small molecules, and cellular markers in applications such as oral cancer, ovarian cancer, prostate cancer, drugs of abuse, cardiac heart disease, and trauma.^18-21^ Previously, we developed the Cardiac ScoreCard system for predicting a spectrum of cardiovascular disease.^22^ This scoring system combines multiple risk factors and biomarker measurements to provide personalized reports for a range of disease indications with diagnostic and prognostic models for cardiac wellness, acute myocardial infarction, and heart failure. The new study described here leverages our past experiences developing clinical decision support tools to efficiently adapt our flexible platform for the development of a prognostic test for COVID-19.

This paper describes the customization of a point-of-care diagnostic tool that is suitable for the measurement of biomarkers that can be used to discriminate between COVID-19 patients that recover vs. those that die from complications of this terrible disease. The work details both the development of a multiparameter protein assay and the diagnostic models that can lend information related to the COVID-19 severity. The model was trained and internally validated using data from 160 hospitalized COVID-19 patients from Wuhan, China^14^ and was evaluated on an external case study of 12 hospitalized patients with a spectrum of COVID-19 disease complications from Shenzhen, China. To our knowledge, this effort is the first quantitative point-of-care diagnostic panel linked to a clinical decision support tool that could be used to predict disease severity for patients suffering from COVID-19 infections. In addition to the new point-of-care diagnostic panel and decision tools, an app is envisioned for immediate release to help clinicians in the next few weeks manage their COVID-19 patients.

## Materials and methods

### Cartridges

The design and fabrication of single-use disposable p-BNC cartridges equipped with a dedicated biohazardous waste reservoir used in this study were published previously.^23^ To summarize, the cartridges comprised an injection-molded fluidic body and laminate capping layers on top and bottom sides. The upper capping layer was patterned with fluidic channels and through-holes. Aluminum blister packs were bonded to the cartridge’s upper DSA (double sided adhesive) layer with 1µm super hydrophobic polyvinylidene fluoride (PVDF) membranes (EMD Millipore, Billerica, MA). Debris filters were made with 3µm Whatman® Nuclepore Track-Etch Membrane (GE Healthcare, Fairfield, CT). A polyethylene terephthalate (PET) capping layer covered the remaining exposed adhesive.

### Instrumentation

While the fully integrated point-of-care instrumentation has been described previously,^23^ for this current study the instrument was configured into a modular fixture for experimentation and assay development. The instrument was manufactured by Open Photonics Inc. (Orlando, FL) and XACTIV Inc. (Fairport, NY). The blister actuator module featured two linear actuators and a motor controller secured to a machined aluminum support framework. Two linear actuators (Haydon Kerk Motion Solutions, Inc., Waterbury, CT) were fitted with force sensitive resistors (400 series, Interlink Electronics, Inc., Westlake Village, CA). The optics module was constructed from threaded lens tubes and adapters (Thorlabs Inc., Newton, NJ) mounted onto a machined aluminum support base. Excitation light was provided by a 490nm LED and T-Cube LED Driver (Thorlabs Inc., Newton, NJ). Optical filters included a 520/15nm BrightLine® single-band bandpass emission filter (Semrock, Inc., Rochester, New York), a 466/40nm excitation filter, and a 506nm dichroic mirror (Edmund Optics, Barrington, NJ). Images were captured on a Grasshopper®3 camera with a Sony IMX174 CMOS sensor (Point Grey Research, Inc., Richmond, British Columbia, Canada). Control software and user interface was developed in MATLAB® 2014a (Natick, MA).

### Immunoassay

A multiplex immunoassay was developed for a subset of the proposed biomarkers to demonstrate proof of concept for the COVID-19 disease severity panel. Spherical agarose sensor beads (2% cross-linked) were synthesized using methods previously reported.^24^ Beads were then sorted into a narrow size distribution (280 ± 10 μm) using test sieves, cross-linked, and glyoxal activated. Activated beads were then functionalized with analyte-specific capturing antibodies using reductive amination with 50mM sodium cyanoborohydride followed by deactivation of unreacted sites in 1M tris buffer with 50mM sodium cyanoborohydride.

The cTnI and NT-proBNP antibodies and standards were purchased from HyTest, Ltd., (Turku, Finland). CK-MB, CRP and Goat anti Mouse IgG (H + L) (R-PE) specific antibodies and standards and were acquired from Fitzgerald Industries International (Acton, Massachusetts). MYO-specific antibodies and standards were acquired from Meridian Life Sciences Inc. (Memphis, TN). Mouse monoclonal anti-human antibodies for cTnI, (clone M18 and 560), CK-MB, MYO (clone 7C3), NT-proBNP (clone 15C4), CRP, and goat anti mouse IgG (H + L) (R-PE) antibodies were conjugated to beads sensors for target capture. Alexa Fluor 488 was conjugated to cTnI, (clone 19C7 and 267), CK-MB, MYO (clone 4E2), NT-proBNP (clone 13G12), and CRP antibodies using Alexa Fluor 488 protein labeling kit (Invitrogen, Eugene, Oregon) for target detection using manufacturer specified protocols.

Cartridges were manually populated with bead sensors and conjugate pad reagents. Bead sensors were strategically configured into designated locations within a 4×5 bead support chip for spatial identification. Detection antibodies were spotted onto a 2×15mm glass fiber conjugate pad (EMD Millipore, Billerica, MA) which was inserted into the cartridge. All assays were performed in direct sandwich-type immunoassay format at room temperature. For each assay, the sample was wetted over the sensor array for 15 seconds. The sample was then delivered for 10 minutes at 10µL/min followed by a 15 second wash at 200µL/min. The detecting antibody was eluted from the reagent pad for 1 minute at 100µL/min by flowing PBS through the pad originating from the blister. This was followed by a 5-minute final wash using a ramping flow rate. The total time of the assay was approximately 16 minutes consuming a total volume of 1400µL.

### Image analysis

Images were analyzed using a custom image analysis tool developed with MATLAB as described previously.^23^ The fluorescence response of each bead was expressed as the average pixel intensity for a region of interest limited to the outer 10% of the bead diameter where the specific signal is concentrated. Bead sensors that were optically obstructed by debris or bubbles were excluded from analysis. Likewise, failed assay runs due to leaks were rejected and re-assayed. Curve fitting routines were processed in MATLAB® R2017b.

### Standard curves

Beads were arranged column-by-column in the 4×5 chip. Two mouse-antibody sensitized beads were configured in the upper positions of the far-left column to serve as positive controls which respond to dye conjugated mouse-based antibodies used to visualize the target. Two CRP-sensitized beads were positioned in the lower positions of the far-left column to serve as negative controls. Both positive and negative controls represent internal QA/QC beads where the response parameters can be used as the basis for run rejection in the event of an error. Sensor beads cTnI, CK-MB, MYO, and NT-proBNP were arranged in a 4-fold redundancy in the remaining columns. Once the beads were in place, the silicone coated release liner was removed from the chip, and an optical cover was bonded to the exposed underlying adhesive sealing the analysis chamber. A cocktail of cTnI, CK-MB, MYO, and NT-proBNP standards were prepared in goat serum (Meridian Life Sciences) at concentrations of 500, 100, 20, 4, 0.8, 0.16, and 0.032 ng/mL. Standards solutions were processed on the p-BNC assay system in triplicate, and their responses were determined. Five matrix blank samples were also processed to determine the variation of the blank response. The upper end of the assay range was determined as the highest concentration achievable without saturating the sensor beads.

### Model Development and Statistical Analysis

This study involves the development of a COVID-19 Severity Score using similar methods as described previously.^22^ Biomarker data from 160 hospitalized COVID-19 patients were derived from a recent study in Wuhan, China.^14^ Patients were assigned to two outcomes: patients who were discharged (n=117) and patients that died (n=43). A lasso logistic regression model for COVID-19 was trained using the following as predictors: age, sex, cTnI, CRP, PCT, and MYO. The maximum biomarker values across all time points were extracted for each patient and log transformed. Then, all data were standardized with zero mean and unit variance. Missing data were imputed using the multivariate imputation by chained equations (MICE) algorithm in statistical software R.^25^ Ten imputations were generated using predictive mean matching and logistic regression imputation models for numeric and categorical data, respectively. The data were partitioned using stratified 5-fold cross-validation to preserve the relative proportions of outcomes in each fold. Model training and selection were performed on each of the 10 imputation datasets. Models were selected for the penalty parameter corresponding to one standard error above the minimum deviance for additional shrinkage. Model performance was documented in terms of AUC and median (interquartile range [IQR]) COVID-19 Severity Scores of patients that died versus those that recovered using pooled estimates. COVID-19 Severity Scores from 5-fold cross-validation, and pooled imputed data sets informed boxplots and scatterplots. Biomarker values and COVID-19 Scores were compared for discharged patients vs. those that died using Wilcoxon rank sum test. Age was compared using an independent *t*-test. Proportions were compared using the Chi-squared test.^26, 27^ Two-sided tests were considered statistically significant at the 0.05 level.

We externally validated the COVID-19 Severity Score on data from a case study of 12 hospitalized COVID-19 patients from Shenzhen, China.^28^ Results were presented in a scatter/box plot of COVID-19 Severity Scores on three groups of patients defined as follows: moderate (patients whose only complication was pneumonia), severe (patients with both pneumonia and ARDS), and critical (patients with one or more of severe ARDS, respiratory failure, cardiac failure, or shock).

## Results and discussion

The biomarker profiles for COVID-19 patients change over the timeline of infection. Therefore, there is a need for a series of diagnostic tests that collectively cover/monitor the entire timeline of infection. Here, three tests are relevant. The first is a molecular diagnostic that tests for the virus itself or part of the same. These tests include RT-PCR or immunological tests that are specific for a component of the virus such as the coronavirus spike glycoprotein.^29^ Both assay modalities lend information on the amount of virus present during the initial stages of infection (i.e., days 2 to 20) but lack accurate quantitation information as the samples are often collected from a nasal swab where the sample volume is ill-defined. After this initial infection phase, the virus itself becomes suppressed due to the activation of the humoral response of the host that involves production of anti-virus specific antibodies.

The second relevant diagnostic test involves detecting this antibody response as an indicator of exposure and subsequent immune response to the virus. The humoral immune response usually begins with the production of IgM antibodies. IgM antibodies tend to have low affinity since they are produced before B cells undergo somatic hypermutation; however, IgM is pentameric in structure, making it an effective activator of the complement system which is important in controlling infections of the bloodstream. The monomeric IgG dominates the later phases of the antibody response.^30^ A test for coronavirus exposure and immune response uses viral antigen to detect these antibodies in the bloodstream.

Throughout this timeline it would benefit care decisions and planning for resource allocation to identify those high-risk patients with underlying, ongoing, or past medical conditions. The sooner these patients are identified, the better is their prognosis through stabilizing measures and close monitoring. As highlighted in the Introduction, one of the major diagnostic gaps and the focus of this paper is determining COVID-19 disease severity, which is the third relevant diagnostic test associated with COVID-19. Initial reports suggest that COVID-19 is associated with severe disease that requires intensive care in approximately 5% of proven infections.^8^ A report from the Chinese Center for Disease Control and Prevention stated that the case fatality rate was higher for those with cardiovascular disease (10.5%), diabetes (7.3%), chronic respiratory disease (6.3%), hypertension (6.0%), and cancer (5.6%). Growing evidence suggests that COVID-19 interacts with the cardiovascular system on multiple levels with increased morbidity and mortality in those with underlying cardiovascular conditions.^31^ Further, evidence of myocardial injury has been observed at higher rates in those that died.^31^ In a study of 187 patients, 7.6% of patients (8 of 105) with normal cardiac troponin T levels and without cardiovascular disease died versus 69.4% of patients (25 of 36) with both elevated cTnT and cardiovascular disease.^14^ The underlying health of the patient has a strong association with COVID-19 outcomes and must be included in clinical decision support tools for determining disease severity.

With this perspective in mind, development of a portable assay system suitable for COVID-19 disease severity would be extremely important in the coming weeks and months as the global pandemic moves forward. Given the broad spectrum of disease severity and rapid clinical decline of patients who develop pneumonia and/or cardiac injury, a point-of-care assay and decision support system could improve triage of patients—and eventually outcomes—for those who need more immediate and aggressive care. Incorporating the calculation of the COVID-19 Severity Score into electronic health records (EHR) would provide health providers with actionable information at an early stage so resources can be focused on patients who are expected to be most severely affected. The measurements of the proteins included in the score can either be provided by EHR integration of the point-of-care biosensor system described here or collected from multiple separate test platforms. Most widely used EHRs support the construction of custom-made decision support systems allowing a fast implementation of the COVID-19 Severity Score based on currently available methods for measuring the proteins used for calculating the score. The EHR integration of the point-of-care biosensor system can follow later once it is validated for this indication. This stepwise approach will allow a fast deployment of the COVID-19 Severity Score followed by an increased testing throughput through the implementation of the point-of-care biosensor system. This will allow better triage of patients and allow scarce healthcare resources to be focused on the patients most at risk for developing severe symptoms.

The p-BNC, a point-of-care biosensor system with the capacity to learn, is adapted here for the application of COVID-19 disease severity. **Figure 1** highlights the key diagnostic infrastructure required to complete the integrated biomarker assays as needed to establish the COVID-19 Severity Score. From a small amount of patient sample (∼100 μL serum), the cartridge and instrument perform a multistep assay sequence to ‘digitize biology’ by converting fluorescence immunoassay signal into biomarker concentrations. Statistical learning algorithms trained on data of biomarker studies predict a spectrum of disease. The result is a single value score which can be displayed to patients and providers in a mobile health app or directly on the instrumentation completing the test. Previously, we published a general framework for implementing a point-of-care based clinical decision support system.^17, 22^ Here, we have adapted these methods to the task of predicting mortality in patients with COVID-19. It should be emphasized that while the integrated testing and COVID-19 Severity Score reporting here articulated represent what is arguably the most efficient delivery mode, the scaling and regulatory approval for this test ecosystem will take several months to complete. With the imminent arrival of the peak of the COVID-19 pandemic, it is important to emphasize that the COVID-19 Severity Score can be generated immediately using biomarker measurements collected from multiple separate test platforms. We anticipate this contribution could have an immediate impact on COVID-19 patient management, and we plan to promptly distribute the COVID-19 Severity Score capabilities following additional model refinement and validation.

**Figure 1.**
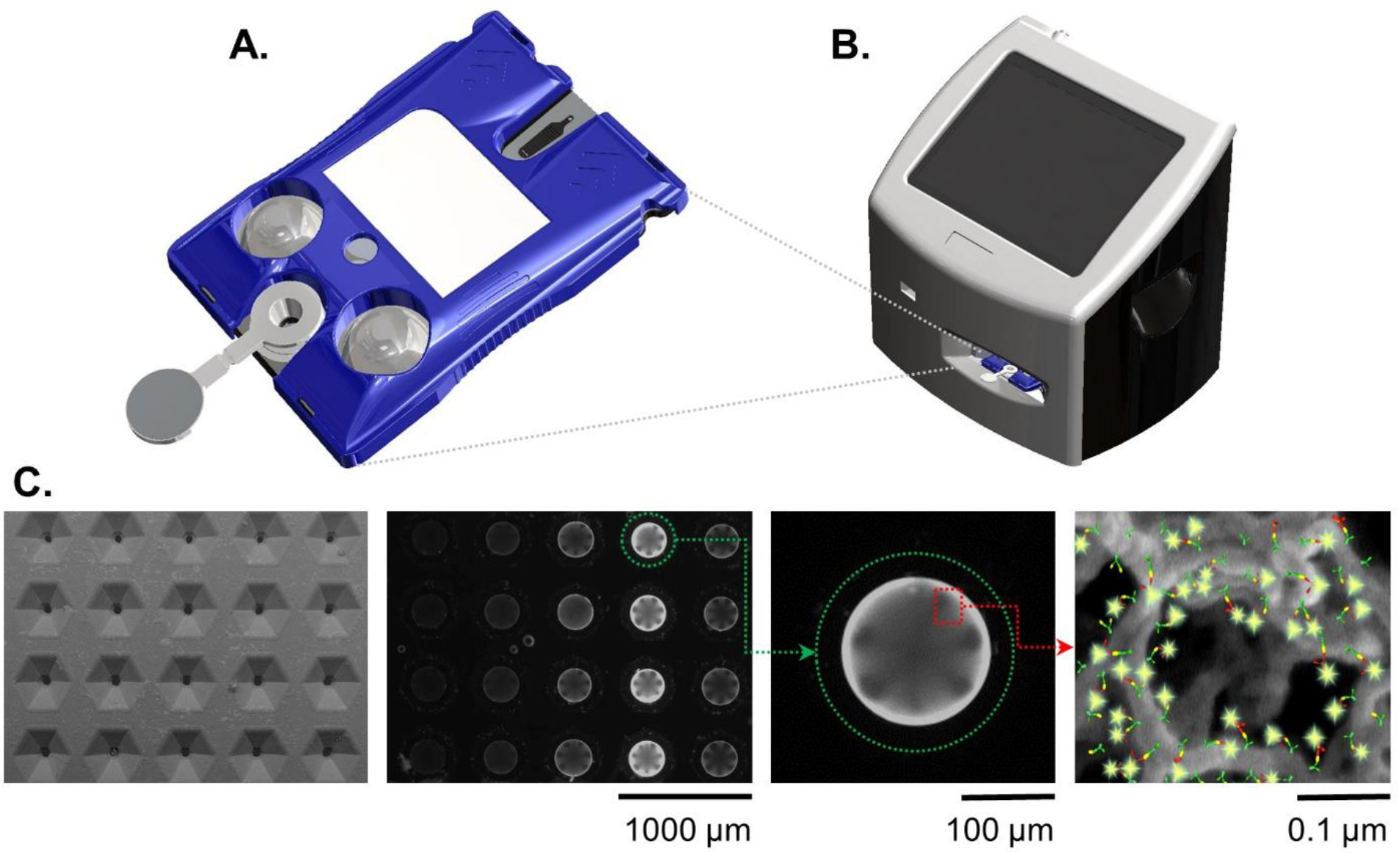
The p-BNC assay system consists of a disposable cartridge (A) and a portable instrument (B). The instrument facilitates fluid motivation inside the cartridge by crushing the fluid filled blister packs on the cartridge surface and reads the resulting optical fluorescent signal generated on bead sensors (C) (from left to right: SEM image of the cartridge’s bead array chip; fluorescent photomicrograph of the bead sensors; an agarose bead sensor with immunofluorescent signal; illustration of a sandwich immunoassay on agarose bead fibers).

Selection of the biomarkers targeted for the development of this COVID-19 Severity Score was based on the following process/criteria and summarized in **Table 1**. Biomarkers were identified as relevant to complications associated with COVID-19 including those associated with acute inflammation/infection (CRP) and various stages of the cardiovascular disease such as coronary artery disease (CRP, PCT), acute myocardial infarction (cTnI, myoglobin), and congestive heart failure (NT-pro BNP, D-dimer). The biomarker PCT, an aid in decision making on antibiotic therapy for hospitalized patients or in patients with suspected or confirmed lower respiratory tract infections, including community-acquired pneumonia, acute bronchitis, and acute exacerbations of chronic obstructive pulmonary disease, was also selected as a valuable tool in the COVID-19 pandemic to identify patients at risk for bacterial coinfection and adverse outcomes. Importantly, all the selected biomarkers have reportedly been shown to exhibit significant differences in their levels in COVID-19 patients that recover vs. those that die.

**Table 1.** COVID-19 disease panels targeted for the applications of disease severity and community surveillance. While this current study presents the framework of a COVID-19 Severity Score for disease severity, future work will involve developing a rapid test of coronavirus exposure for surveillance applications using the same programmable diagnostic platform here featured.

| Panel | Analytes | Comments |
| --- | --- | --- |
| Severity | CRP | Evidence of infection or inflammation |
|  | PCT | Inflammatory marker; mortality indicator |
|  | CK-MB | Elevated in COVID-19 patients, myocardial infarction |
|  | cTnI | Myocardial infarction, heart failure |
|  | D-dimer | Thrombotic events, myocardial infarction, heart failure |
|  | Myoglobin | Myocardial infarction, COVID-19-associated rhabdomyolysis |
|  | NT-proBNP | Heart failure |
| Surveillance | Spike protein | Viral antigen |
|  | IgG | Most abundant type of antibody |
|  | IgM | First antibody made to fight a new infection |
|  | SIgA | Secretory Immunoglobulin A (SIgA) is the main immunoglobulin found in salivary glands and plays a key role in protecting from invading pathogens |

Although the p-BNC is designed to accommodate both soluble and cellular targets using either bead or membrane-based assay configurations, respectively, we opted to solely focus on soluble protein biomarkers. Further, we restricted biomarker choices to those that have complementary concentration ranges and those that are stable allowing for their simultaneous measurement. Though lymphocytes and cytokines have been associated with COVID-19 mortality, neither of these two classes of analytes were selected because of their incompatibility with these selection criteria.

The complementary COVID-19 assay panels for disease severity index (described here) and surveillance panel (to be featured in future publications) are shown along with their relevant immunoschematics in **Figure 2**. Briefly, bead-based tests for the severity index panel targets the simultaneous measurement of six designated proteins, all compatible for multiplexed detection. In this direct sandwich immunoassay involving six matched pairs of capture/detection antibodies, all six biomarkers are first captured by their corresponding beads and then specifically detected via their matched Alexa Fluor 488-conjugated detection antibodies presented to the bead array. During the development of these fully quantitative assays, control experiments are conducted to ensure that there is no crosstalk (interference) between each of the assays.

**Figure 2.**
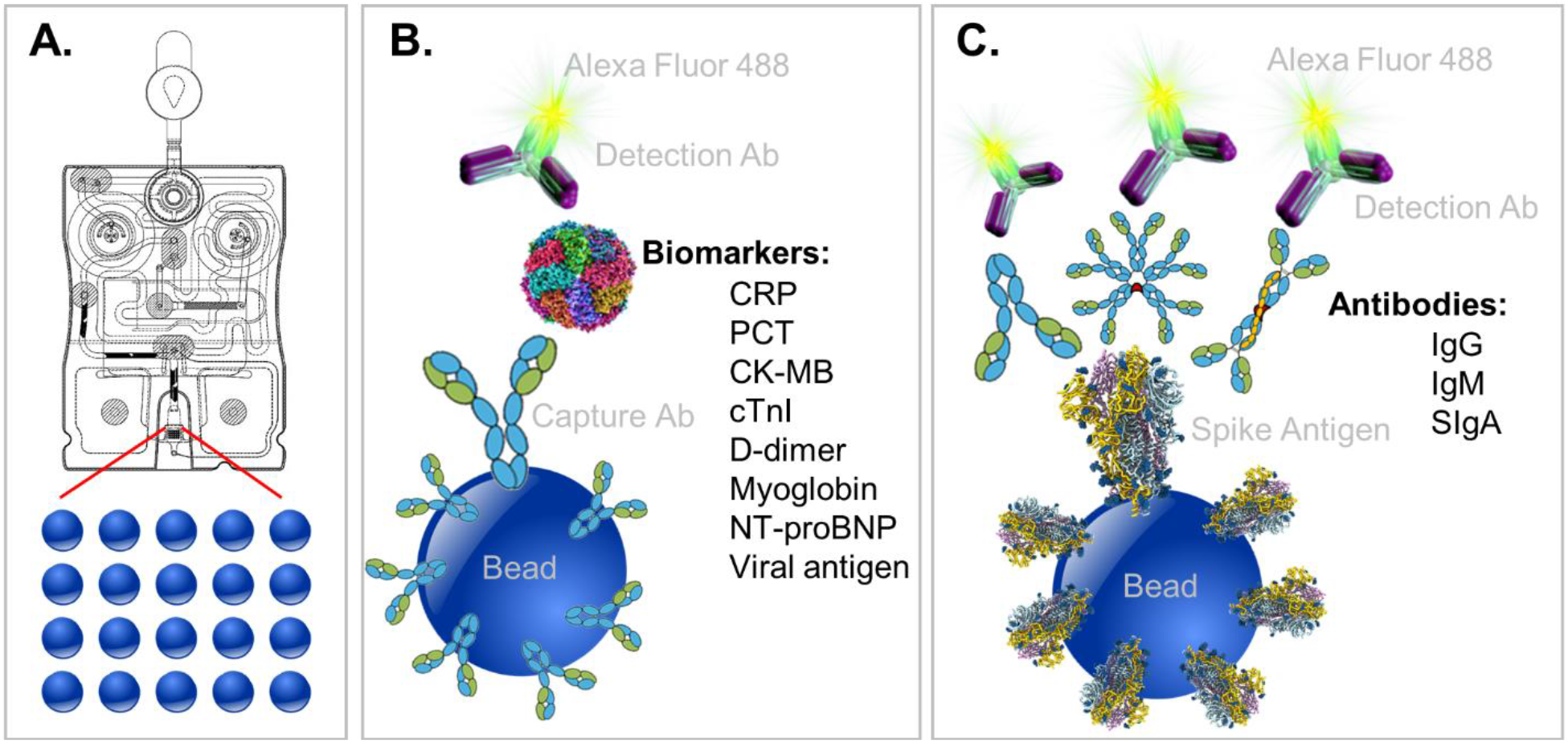
Programmable cartridge for COVID-19 diagnostics. The p-BNC cartridge features 20 spatially programmable bead sensors (A) that can be customized for a multitude of applications. Here, two panels are detailed for COVID-19: a disease severity panel as featured in the work (B) and a community exposure / surveillance panel as will be described in future efforts (C).

A multiplex immunoassay was developed for a subset of the proposed biomarkers to demonstrate proof of concept for the COVID-19 disease severity panel. The p-BNC platform can perform powerful and quantitative multiplexed measurements over an extended range. Calibration curves are necessary to quantitate the concentration of molecular targets in solution which are critical inputs to the diagnostic algorithm. **Figure 3** demonstrates this capability with four simultaneously generated calibration curves for cTnI, CK-MB, MYO, and NT-proBNP that cover a concentration range from 0.032 to 500 ng/mL. Error bars indicate bead-to-bead precision with four redundant beads measured per sensor class. As shown, the response data for each biomarker exhibits an excellent fit to a five-parameter logistic regression. As part of the multiplexed assay development effort, specificity was confirmed for the four-plex panel, as shown in inset images on **Figure 3**. Here, a single antigen standard at high concentration (1000ng/mL) was processed against a cartridge configured for multiplexed detection. As expected, monoclonal antibodies are highly specific for their target antigen, where high doses of each single antigen generated minimal cross-reactivity on non-target sensors. Although this work represents a subset of the full COVID-19 panel, the cartridge facilitates multiplexing of up to 20 different biomarkers and can be easily expanded to accommodate the panel and test validity controls. We anticipate that one or more of the selected six biomarkers may be dropped as additional COVID-19 clinical data are used to optimize the final COVID-19 Severity Score due to redundancy of patient discrimination information afforded by these biomarkers.

**Figure 3.**
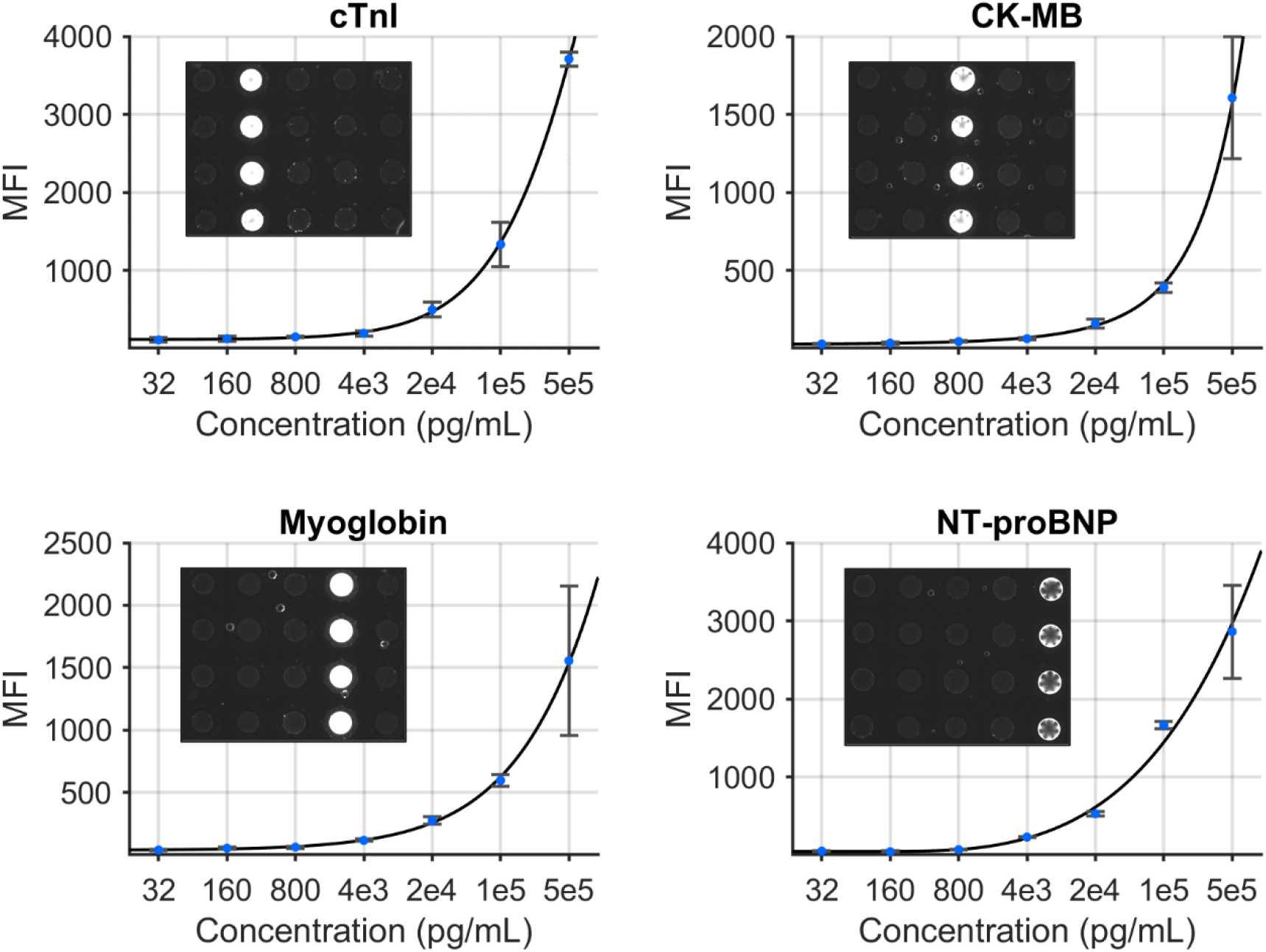
Standard curves generated for a COVID-19 disease severity biomarker panel including cTnI, CK-MB, myoglobin, and NT-proBNP.

Having identified a method to measure strategic biomarkers in a multiplexed panel, this next step involves the translation of these test values alongside key clinical metrics into information relevant to COVID-19 disease severity. A COVID-19 Disease Severity model was developed based on data from 160 hospitalized patients from Wuhan, China.^14^ Here, 160 patients with hypertension were admitted to the hospital for COVID-19 in which 117 were eventually discharged and 43 died. **Table 3** summarizes the patient characteristics and lab values for both patient groups. Interestingly, males accounted for 70% of the deaths vs. 44% of the discharged patients. This study finds significantly higher levels of biomarkers (cTnI, CK-MB, MYO, CRP, and PCT) in patients that died vs. those that were discharged. Likewise, age was a statistically significant factor with mean (SD) of 63 (13) and 73 (8) in the “discharged” and “died” groups, respectively.

**Table 2.** COVID-19 biomarkers from the literature. Values are presented as median (IQR), mean (standard deviation), n (%), and AUC (95% CI).

| Source | COVID-19 Patients | Biomarkers | Case | Noncase |
| --- | --- | --- | --- | --- |
| Huang et al.<sup>6</sup> | ICU care (n=13) vs. No ICU care (n=28) | cTnI, pg/mL | 3.3 (3.0–163.0) | 3.5 (0.7–5.4) |
|  |  | D-dimer, mg/L | 2.4 (0.6–14.4) | 0.5 (0.3–0.8) |
|  |  | PCT, ng/mL | 0.1 (0.1–0.4) | 0.1 (0.1–0.1) |
| Wang et al.<sup>5</sup> | ICU (n=36) vs. Non-ICU (n=102) | cTnI, pg/mL | 11.0 (5.6–26.4) | 5.1 (2.1–9.8) |
|  |  | D-dimer, mg/L | 414 (191–1324) | 166 (101–285) |
|  |  | CK-MB, U/L | 18 (12–35) | 13 (10–14) |
| | | PCT ≥ 0.05 ng/mL | 27 (75.0) | 22 (21.6%) |
| Ruan et al.<sup>35</sup> | Died (n=68) vs. Discharged (n=82) | cTnI, pg/mL | 30.3 (151.1) | 3.5 (6.2) |
|  |  | Myoglobin, ng/mL | 258.9 (307.6) | 77.7 (136.1) |
|  |  | CRP, mg/L | 126.6 (106.3) | 34.1 (54.5) |
| Zhang et al.<sup>10</sup> | Severe (n=58) vs. Nonsevere (n=82) | D-dimer, ug/mL | 0.4 (0.2–2.4) | 0.2 (0.1–0.3) |
|  |  | CRP, mg/L | 47.6 (20.6–87.1) | 28.7 (9.5–52.1) |
|  |  | PCT, ng/mL | 0.1 (0.06–0.3) | 0.05 (0.03–0.1) |
| Guo et al.<sup>14</sup> | Cardiac injury (n=52) vs. No cardiac injury (n=135) | D-dimer, ug/mL | 3.85 (0.51–25.58) | 0.29 (0.17–0.60) |
|  |  | CRP, mg/dL | 8.55 (4.87–15.17) | 3.13 (1.24–5.75) |
|  |  | PCT, ng/mL | 0.21 (0.11–0.45) | 0.05 (0.04–0.11) |
|  |  | CK-MB, ng/mL | 3.34 (2.11–5.80) | 0.81 (0.54–1.38) |
|  |  | Myoglobin, ug/L | 128.7 (65.8–206.9) | 27.2 (21.0–49.8) |
|  |  | NT-proBNP, pg/mL | 817.4 (336.0–1944.0) | 141.4 (39.3–303.6) |
| Chen et al.<sup>2</sup> | COVID-19 patients (n=99) | D-dimer, ug/mL | 0.9 (0.5–2.8) | NA |
|  |  | PCT, ng/mL | 0.5 (1.1) | NA |
|  |  | CRP, mg/L | 51.4 (41.8) | NA |
| Bai et al.<sup>9</sup> | AUCs for Died (n=36) vs. Recovered (n=91) | cTnI, ng/mL | 0.939 (0.896–0.982) | NA |
|  |  | CRP, mg/L | 0.870 (0.801–0.939) | NA |
|  |  | PCT, ug/L | 0.900 (0.824–0.975) | NA |
|  |  | D-dimer, ug/L | 0.866 (0.785–0.947) | NA |

**Table 3.** Summary of patient characteristics and lab values. Data are presented as median (IQR), number (%), mean (SD).

|  | Discharged | Died | p-value |
| --- | --- | --- | --- |
| Patients | 117 | 43 | NA |
| Age, y | 63 (13) | 73 (8) | < 0.001 |
| Sex, male | 52 (44) | 30 (70) | 0.023 |
| cTnI, pg/mL | 5.40 (1.65–8.05) | 121.10 (50.85–306.65) | < 0.001 |
| CK-MB, ng/mL | 4.25 (1.10–11.25) | 5.31 (2.29–18.26) | 0.011 |
| MYO, ng/mL | 45.35 (27.00–78.30) | 177.80 (92.65–896.00) | < 0.001 |
| CRP, mg/L | 18.50 (6.92–63.28) | 140.30 (84.75–248.23) | < 0.001 |
| PCT, ng/mL | 0.05 (0.05–0.11) | 0.55 (0.18–1.46) | < 0.001 |

A COVID-19 Severity Score was trained and internally validated based on a subset of the targeted biomarkers (cTnI, PCT, MYO, and CRP), age, and sex. The disease discrimination potential is displayed in **Figure 4**. For this analysis, COVID-19 Severity Scores are shown for two patient groups, those patients that recovered vs. those that passed away from the complications. The COVID-19 Severity Score is the lasso logistic regression response from internal validation interpreted as the probability of death. Individual points on the scatterplot represent the COVID-19 Severity Score for one sample with overlaid boxplots representing the COVID-19 Severity Score for the population of patients. Additional model information is included in the Supplemental Materials, including model coefficients (**Figure S1**) and AUC values (**Table S1**). The median (IQR) COVID-19 Severity Scores were significantly higher for those that died vs. those that were discharged (59 [40–83] vs. 9 [6–17], respectively). Patients who underwent any invasive or noninvasive ventilation were at an intermediate risk of death with median (IQR) scores of 17 (10– 39) (**Figure S2**). The AUC (95% CI) of the multivariate COVID-19 Severity Score was 0.94 (0.89– 0.99), demonstrating proof of concept for the clinical decision support tool.

**Figure 4.**
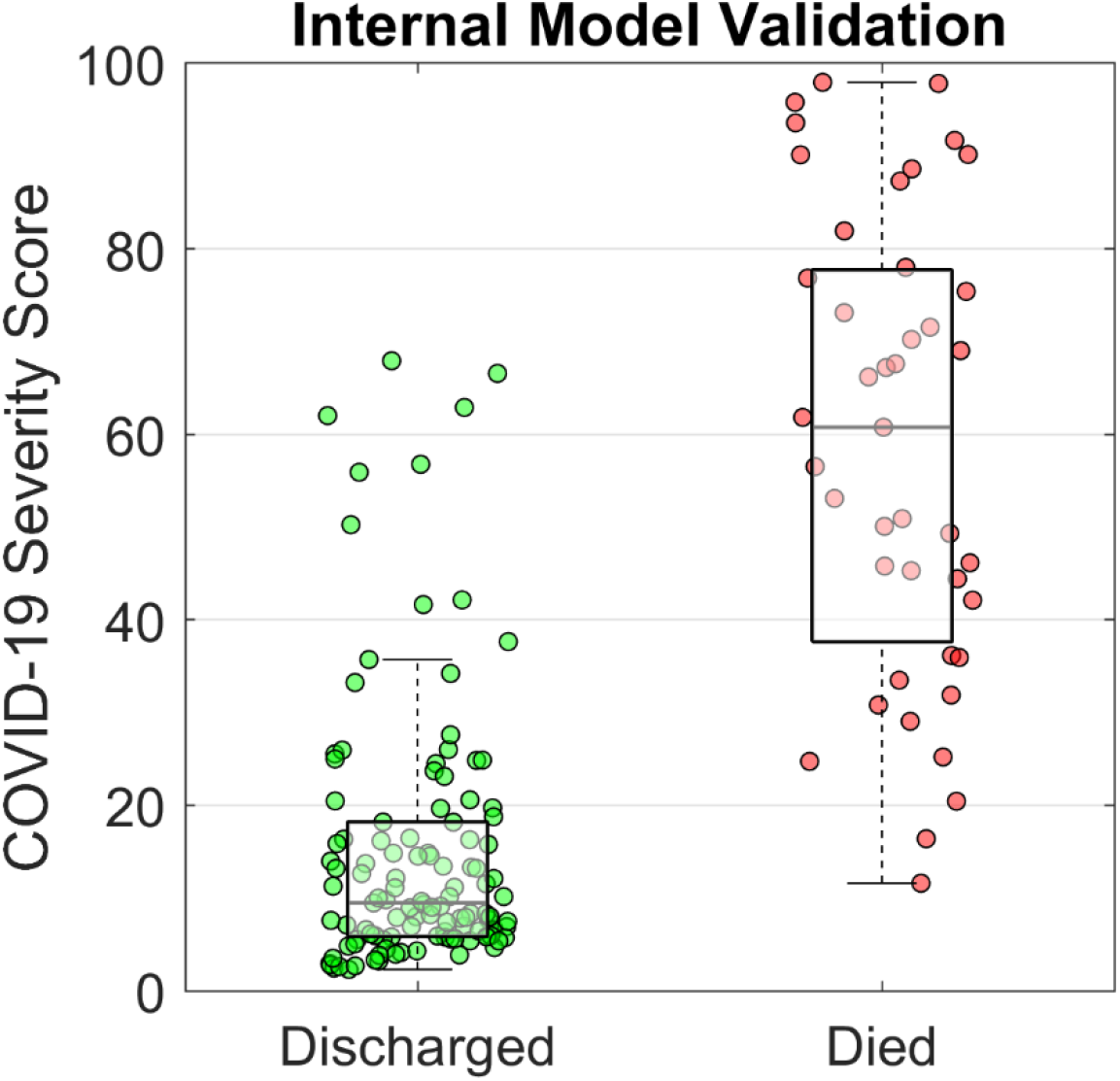
COVID-19 Severity Scores from internal model validation. A model was trained on data from hospitalized COVID-19 patients of which 117 were discharged and 43 died. The COVID-19 Severity Score is a numerical index between 0 and 100 that indicates the probability of COVID-19 mortality. Individual patient scores are represented as scatter dots with overlaid boxplots showing the population distribution.

One limitation of this study was that all patients in the training dataset had hypertension and are, thus, at an elevated risk for cardiovascular events. Since the test panel contains several cardiac biomarkers, it’s possible that these training data could lead to overoptimistic results. However, in addition to cardiac biomarkers, the expanded biomarker panel represents diverse pathophysiology (i.e., indicators of infection, inflammation, mortality, thrombotic events, and rhabdomyolysis) which have the potential to significantly improve generalizability. Plans to evaluate and optimize the COVID-19 Severity Score model on external data are in place. Despite this limitation, the preliminary results demonstrate strong promise for the COVID-19 Severity Score. Reporting these preliminary findings now is critically important given the stage of the pandemic.

Previously we have used the p-BNC platform to develop various wellness and disease severity scores for oral cancer^18, 19, 32^ and cardiac heart disease.^22^ Shown in **Figure 5** is the initial rough scale for the COVID-19 Severity Score which was based on the CDC’s Interim Clinical Guidance for Management of Patients with Confirmed COVID-19.^33^ The continuous scale COVID-19 Severity Score has the potential to assist the identification of patients with severe/critical disease status. For example, most patients (∼80%) with a low COVID-19 Severity Score may be considered at Mild/Moderate risk for developing complications up to mild pneumonia and can be managed at home or in outpatient settings. About 15% of patients may have an elevated COVID-19 Severity Score and would be at risk for Severe disease with complications such as pneumonia, ARDS, sepsis, cardiomyopathy, and others. Approximately 5% of patients may have a high COVID-19 Severity Score that would be considered Critical requiring hospitalization, intensive care, and mechanical ventilation with complications such as respiratory failure, shock, multiorgan failure, and death.

**Figure 5.**
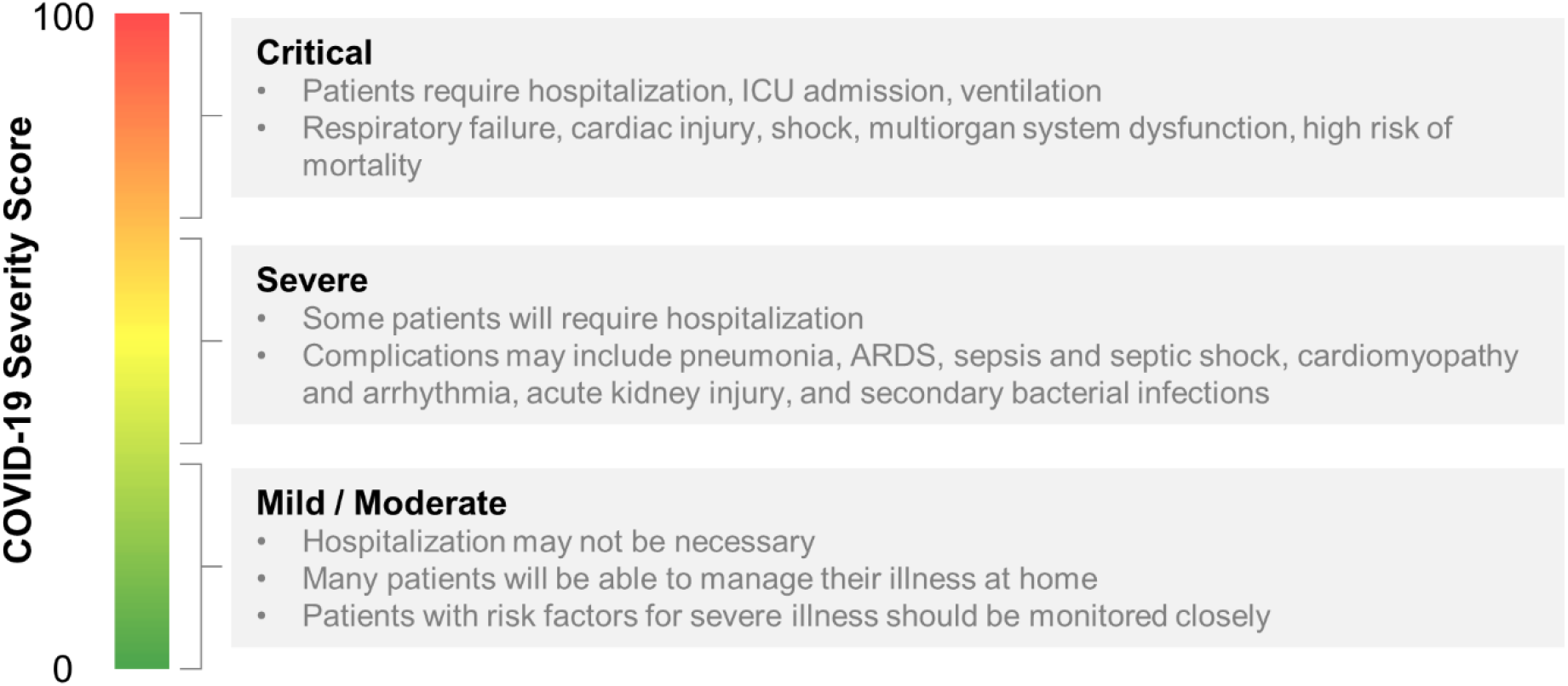
Initial rough scale for COVID-19 Severity Score based on the CDC’s Interim Clinical Guidance for Management of Patients with Confirmed COVID-19.^33^ The continuous scale COVID-19 Severity Score has the potential to assist the identification of patients with severe/critical disease status.

Finally, we evaluated the COVID-19 Severity Score on data from a case study of 12 hospitalized COVID-19 patients.^28^ **Figure 6** presents a scatter/box plot of COVID-19 Severity Scores on three groups of patients. COVID-19 Severity Scores were found to increase with disease severity. Moderate (patients whose only complication was pneumonia), Severe (patients with both pneumonia and ARDS), and Critical (patients with one or more of severe ARDS, respiratory failure, cardiac failure, or shock) groups had median (IQR) COVID-19 Severity Scores of 9 (4–17), 28 (24–36), and 36 (28–83), respectively. Although this analysis evaluates a small sample of patients, these preliminary results show potential for the COVID-19 Severity Score to be calibrated to a disease severity scale. In addition to cross-sectional and population-based comparisons, this COVID-19 Severity Score could also be used for longitudinal monitoring of patients. In this manner, an individual’s time series measurements could be used to track changes in biomarker-based COVID-19 Severity Score over time. Preliminary findings (**Figure S3**) demonstrate that the average trajectories decrease for the “discharged” group increase for the “died” group, suggesting that the COVID-19 Severity Score could provide valuable lead time in discharging patients with low risk earlier while prioritizing care for those at elevated risk of mortality. Future efforts will be used to define various decision cuts points, reference ranges, and change scores to help guide clinical decision making including therapy decisions. Future efforts may also adapt this clinical decision support tool for ARDS resulting from other infectious viral agents such as influenza and varicella-zoster; bacteria such as Mycoplasma, Chlamydia, and Legionella; and parasites such as the malaria causing Plasmodium falciparum.^34^

**Figure 6.**
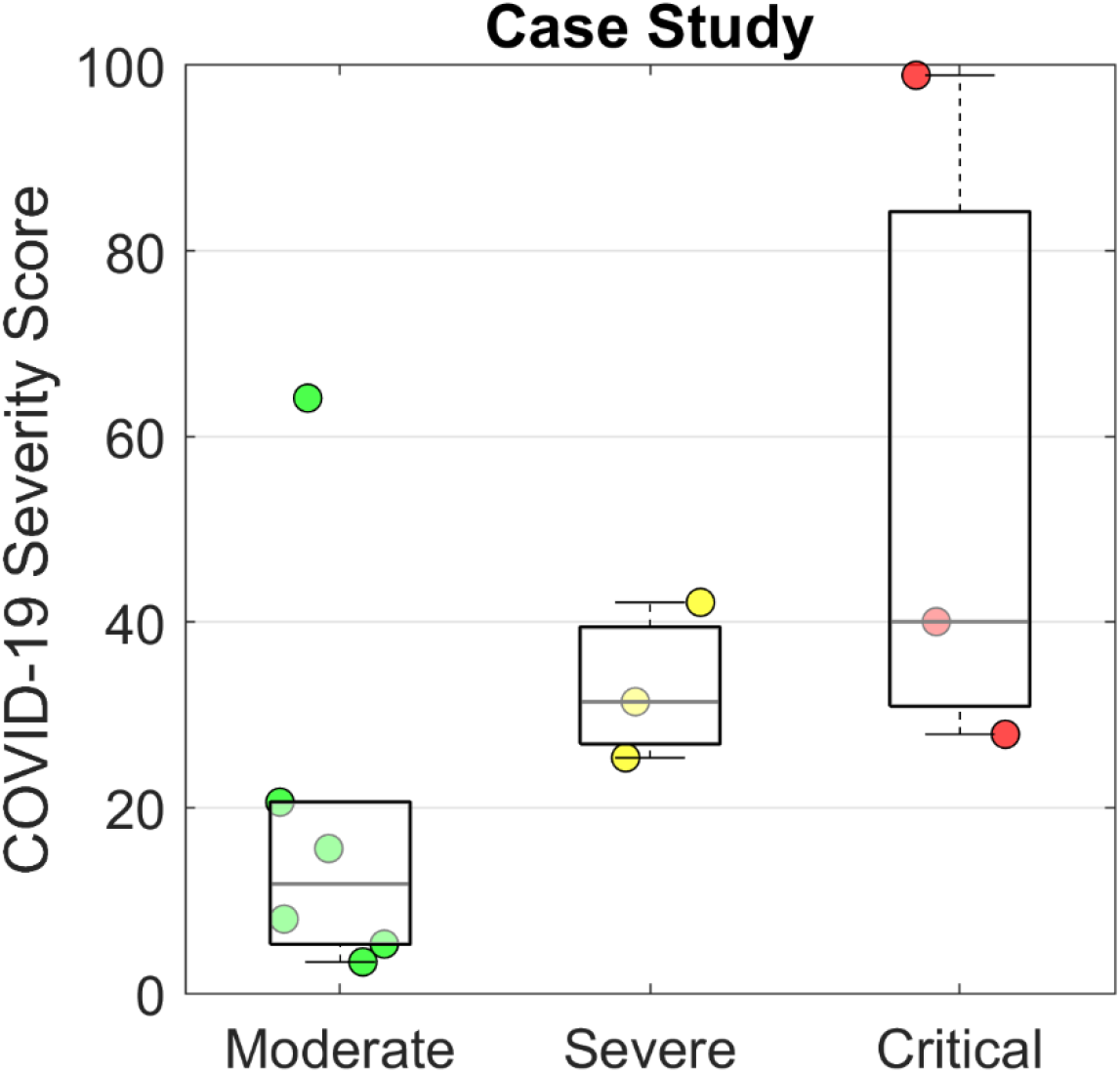
COVID-19 Severity Scores evaluated for a case study of 12 hospitalized patients with COVID-19 from Shenzhen, China.^28^ The Moderate group contained patients whose only complication was pneumonia. The Severe group were patients with pneumonia and ARDS. The Critical group contained patients with one or more of severe ARDS, respiratory failure, cardiac failure, or shock.

## Conclusion

This study establishes the framework for a point-of-care COVID-19 Severity Score and clinical decision support system. Our studies find that the median COVID-19 Severity Score was significantly lower for the group that recovered versus the group that died from COVID-19 complications (60.5 versus 96.6, respectively). The AUC value for the COVID-19 Severity Score was 0.94, demonstrating strong potential for its utility in identifying patients with increased risk of mortality. Plans are now in place to confirm the final selection of biomarkers for an integrated point-of-care COVID-19 Severity Score disease severity test. It is possible that some of the biomarkers may yield redundant information; as such, these redundant biomarkers may be eliminated to create a sparser diagnostic panel with improved generalizability.

These lab-on-a-chip diagnostic capabilities have the potential to yield the first quantitative point-of-care diagnostic panel linked to a clinical decision support tool for predicting mortality from COVID-19. An experienced team and established translation partnerships are both in place to move these systems into real-world practice in a timely manner. Further, the release of an app for immediate impact on COVID-19 patient management in the next few weeks is anticipated. Future work may also involve developing a test on the same platform for population-based COVID-19 community surveillance in clinical settings (ambulances, hospitals, clinics, laboratories) and for public settings that are at risk for community spread (businesses, schools, airports, train stations). The development and distribution of a portable, affordable, widely distributed smart sensor technology with anticipated availability/readiness within months promises to be an important solution for the management of the current coronavirus crisis as well as an adaptable tool to combat future threats of a new virus or biological threat. Likewise, in addition to this COVID-19 Severity Score, a sustaining contribution of this work may be in the development of an ARDS clinical decision support tool for other infectious viral agents, bacteria, and parasites.

## Data Availability

The data that support the findings of this study are available from the corresponding authors, upon reasonable request.

## Acknowledgements

Funding was provided by NIH through the National Institute of Dental and Craniofacial Research (NIH grant no. 3U01DE017793-02S1 and 5U01DE017793-2). The content is solely the responsibility of the authors and does not necessarily represent or reflect views of the NIH, or the Federal Government.

## Supplemental Figures and Tables

**Figure S1.**
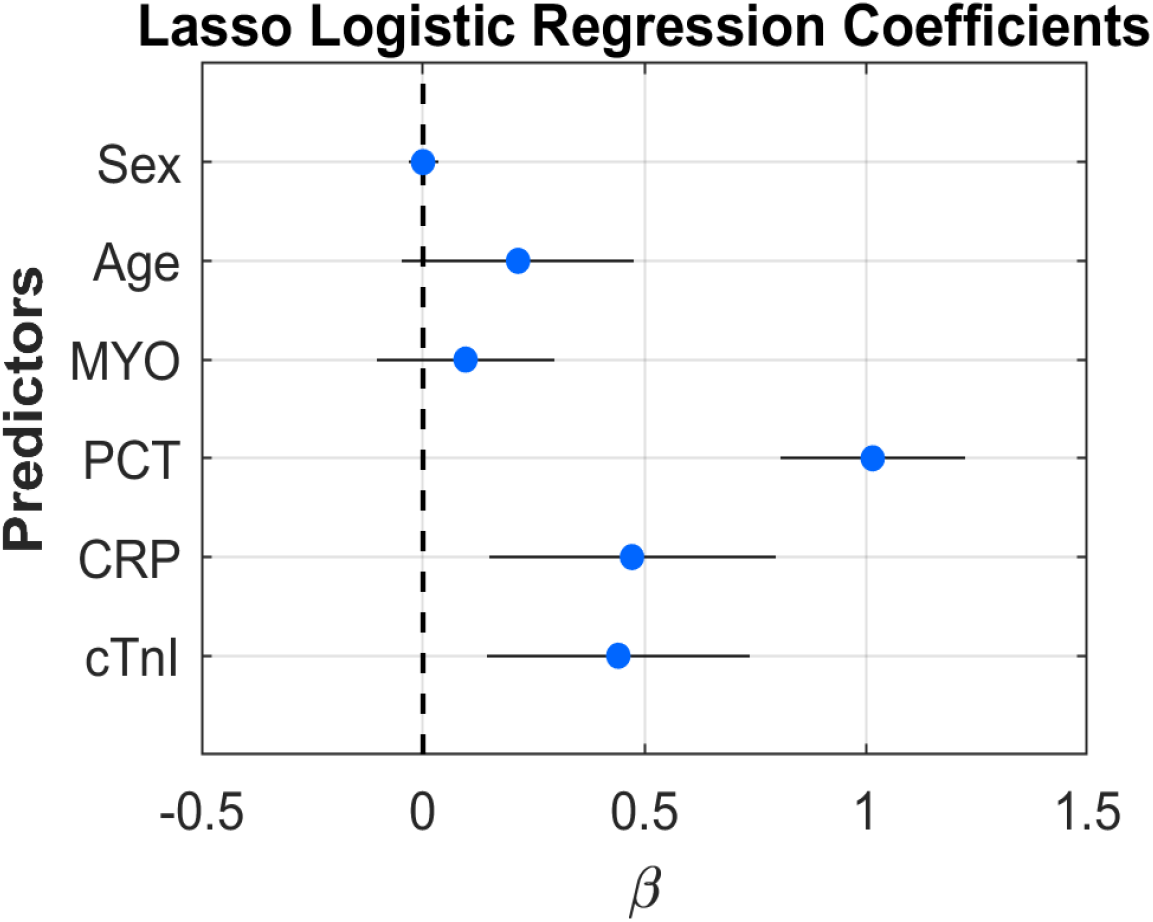
Lasso logistic regression coefficients for the COVID-19 Severity Score model. Coefficients are represented as mean (SD) from 5-fold cross-validation across 10 imputed datasets.

**Figure S2.**
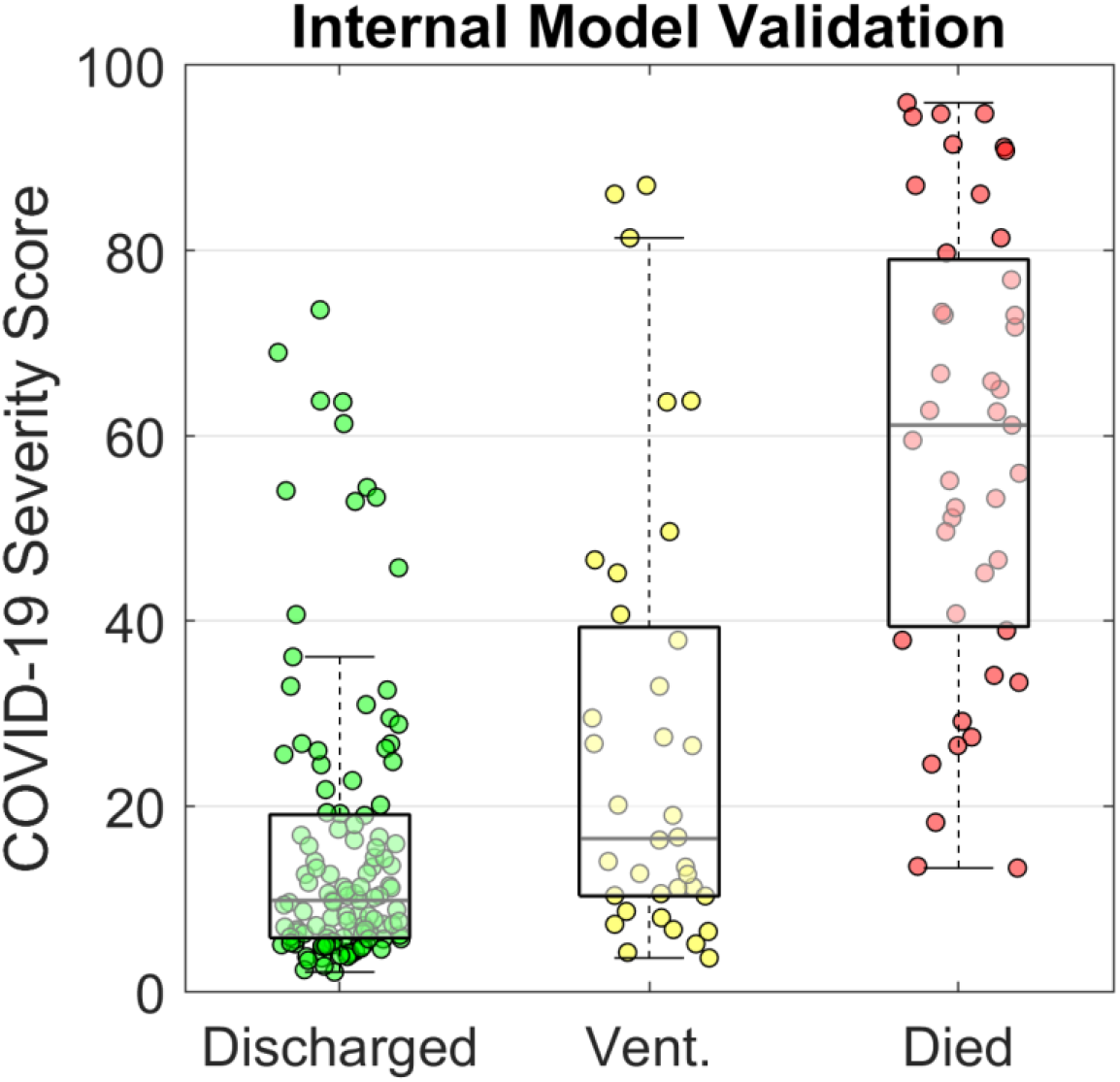
COVID-19 Severity Scores from internal model validation. A model was trained on data from hospitalized COVID-19 patients of which 117 were discharged and 43 died. Out of the total 160 patients, 36 received either invasive or noninvasive ventilation (“Vent.”). The COVID-19 Severity Score is a numerical index between 0 and 100 that indicates the probability of COVID-19 mortality. Individual patient scores are represented as scatter dots with overlaid boxplots showing the population distribution.

**Table S1.**
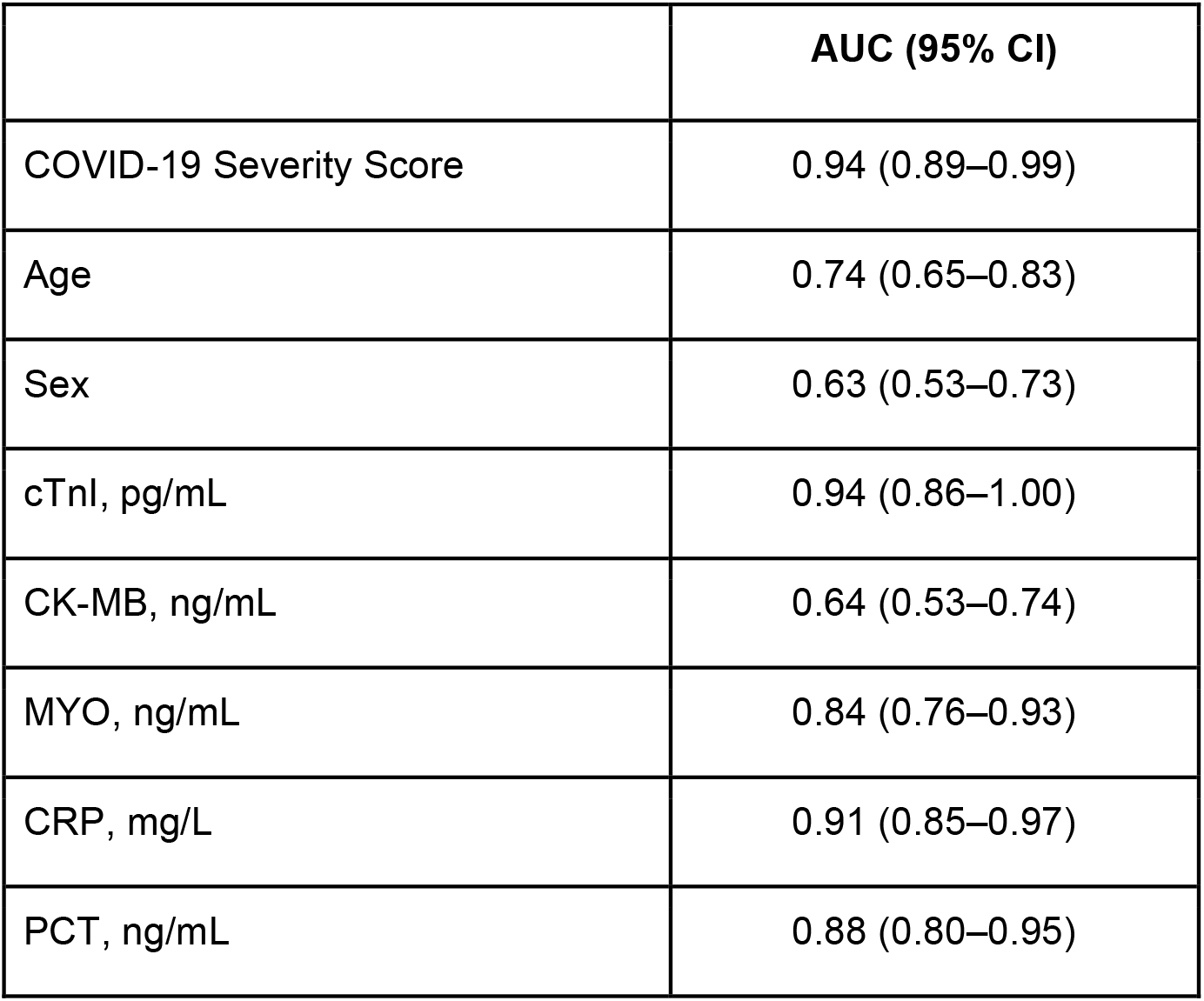
Diagnostic performance of the COVID-19 Severity Score, relevant patient characteristics, and biomarkers.

**Figure S3.**
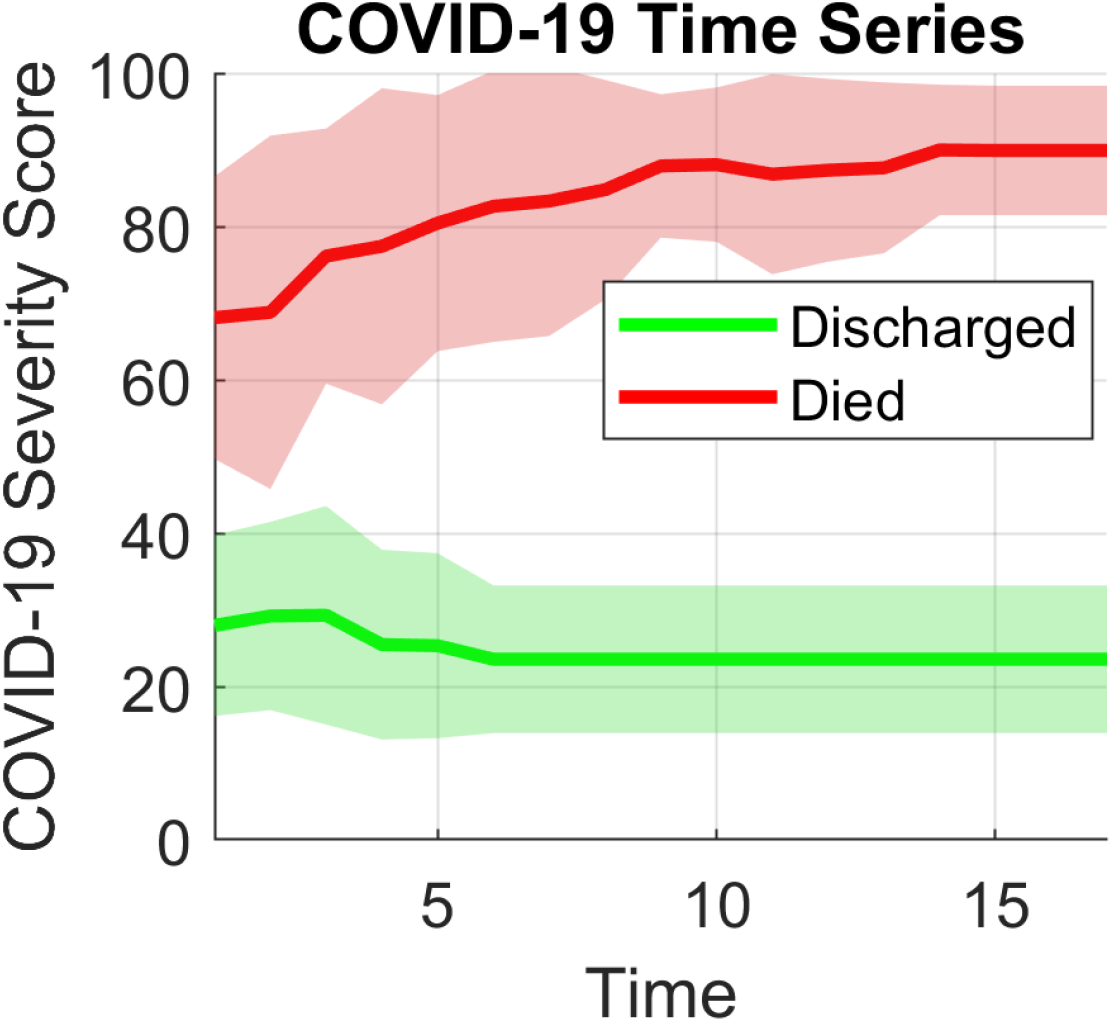
A selection of patients with multiple time course biomarker measurements were analyzed for their COVID-19 Severity Score. Solid lines represent the average scores for all patients in the group, and shaded regions show one standard deviation above and below the mean. The unit of time is arbitrary with 1 being the first measurement after admission and 17 being the last. Missing biomarker values were imputed according to their value at the previous time point.

